# Typologies of activity-related behaviours from childhood to adolescence and their transitions: A longitudinal analysis of the ELSPAC cohort

**DOI:** 10.1101/2024.05.16.24307481

**Authors:** David Janda, Aleš Gába, Ana Maria Contardo Ayala, Anna Timperio, Lenka Andrýsková, Pavel Piler, Lauren Arundell

## Abstract

**Objectives:** To identify typologies of activity-related behaviours in childhood and adolescence and to explore transitions between the identified typologies. Additionally, we aimed to identify demographic indicators associated with the transitions and typology membership.

**Design:** Prospective cohort study

**Setting:** Czech Republic

**Participants:** Individuals involved in the Czech part of the European Longitudinal Study of Pregnancy and Childhood (ELSPAC-CZ) study, aged 11 to 18 years. The study involved more than 563 individuals of which 380 provided complete data the analysis.

**Primary outcome measures:** Time spent outdoors, participation in organised physical activity (PA) and sport activities, time watching television and using personal computer, and total sleep duration. Latent variables defined by the Latent Transition Analysis.

**Results:** Four typologies of activity-related behaviours were identified and labelled to reflect their behavioural profiles: 1) *Actives* (high outdoor time and organised PA and sport participation, low screen time, optimal sleep duration), 2) *Active screeners* (median outdoor time, high organised PA and sport participation, high screen time, optimal sleep duration), 3) *Poor sleepers* (average outdoor time and organised PA and sport participation, low screen time, not meeting sleep guidelines), and 4) *Averages* (average duration of all behaviours and optimal sleep duration). A major shift in typology membership from 11 to 18 years was observed with a decreasing proportion of individuals in typologies characterised by a high proportion of outdoor time and participation in organised PA and sport activities (i.e. *Actives*; *Active screeners*). A high proportion of individuals also transitioned to the typology with poor sleeping habits (i.e. *Poor sleepers*). Sex and maternal education were associated with the typology membership and transition probabilities (*p* < 0.05).

**Conclusions:** Our findings highlight the importance of implementing lifestyle interventions in childhood to prevent possible sleep disorders and low physical activity later in life.

**Strengths and limitations of this study:**

- The longitudinal design allows an investigation of unique activity-related behaviour patterns across the transition from childhood to adolescents.
- Utilising various activity-related behaviours (e.g. outdoor time or screen time) provide a comprehensive understanding of the lifestyle of children and adolescents.
- Data were collected from a specific geographic region from the Central Europe, which may limit the generalisability to broader population.

## Introduction

Childhood and adolescence are critical stages of human development, where rapid physical, mental, and social changes occur [1]. During these formative years, individuals engage in various lifestyle behaviours and form habits contributing to their overall physical and mental health [2, 3]. Previous evidence has shown that healthy lifestyle habits, characterised by adequate physical activity (PA) [4], reduced sedentary behaviour (SB) [5], and optimal sleep [6] developed during childhood and adolescence, can persist into adulthood. Therefore, this period of life offers a valuable opportunity to promote a healthy lifestyle and establish beneficial routines, potentially leading to a positive long-term health trajectory.

Previous longitudinal studies have shown a decline in PA levels and sleep duration, and an increase in SB as children progress into adolescence [7-9]. Shifting towards an unhealthy lifestyle can affect both physical and mental health and might result in detrimental health conditions with long-term health consequences [10]. Therefore, understanding the lifestyle patterns (typologies) and the timing of changes in activity-related behaviours that occur in childhood and adolescence is essential for promoting lifelong healthy habits and designing effective interventions tailored to the specific developmental stages of life.

Most of the existing research concentrates on PA, SB, and sleep, limiting our understanding of the role of other activity-related behaviours in health and disease prevention. Consequently, this narrow scope might overlook some specific domains of lifestyle behaviours that might have a substantial impact on overall health and serve as potential targets for intervention strategies. For example, the available evidence highlights the protective role of outdoor time in reducing the risk of diabetes and high blood pressure [11]. On the other hand, excessive screen time, a specific domain of SB, was found to be associated with elevated risk of obesity and depressive symptoms in children and adolescents [12]. However, how these specific activity-related behaviours contribute to forming distinct typologies is unknown.

The systematic review conducted by Parker et al. [13] reflects an intensive effort that has been devoted to identifying typologies of activity-related behaviours in children and adolescents in the past two decades. However, the majority of published studies were cross-sectional, limiting their ability to investigate how these typologies change over time. Moreover, longitudinal designs examining transitions between typologies have primarily focused on the transitions over short periods (e.g. over 3 years between 6 and 9 years of age) [14], and there has been limited exploration of typology changes during critical stages such as the transition from elementary to high school. Although there is a wealth of research on individual activity-related behaviours during childhood and adolescence, there remains a lack of clarity regarding how additional activity-related behaviours (e.g., outdoor time, screen time) contribute to the formation of typologies, and how individuals transition between these typologies over childhood to adolescence. Furthermore, understanding determinants of changes in typologies over time, as in cross-sectional examples [15, 16], is also important to help identify targets for behavioural interventions.

To address this evidence gap, the present study aims to identify typologies of activity-related behaviours in childhood and adolescence, explore transitions between the identified typologies during these critical developmental periods, and identify demographic indicators of the transitions and typology membership.

## Methods

The present study utilised longitudinal questionnaire data from participants residing in the Czech Republic involved in the European Longitudinal Study of Pregnancy and Childhood (ELSPAC-CZ) study. The detailed study protocol and the cohort description are provided elsewhere [17]. Briefly, the ELSPAC-CZ study collected perinatal information on 5151 mother-child pairs from maternal self-report questionnaires in 1991–1992 and followed the children’s development up to the age of 19 years (*n* = 563). Our analysis specifically utilised self-reported data on activity-related behaviours collected when participants were 11, 15, and 18 years old. These ages were chosen as they represent critical transition periods in childhood and adolescence. The final analytic sample consisted of participants who provided complete data of interest in all three time points (*n* = 380).

Informed consent was obtained from all study participants and their parents or legal guardians prior to completing each questionnaire. All methods were performed in accordance with the relevant guidelines and regulations of the Declaration of Helsinki and the procedures were anonymised. The study was approved by the ELSPAC-CZ Law and Ethics Committee and local research ethics committees. The secondary use of all ELSPAC-CZ study data was approved by the (C)ELSPAC Ethics Committee (Ref. No. ELSPAC/EK/1/2014, date 09/17/2014).

### Assessment of activity-related behaviours

Data on activity-related behaviours were collected through a tailored questionnaires which queried participants about the sleep duration, time spent in PA, outdoor time, and in front of screens. Participants were asked to report their time spent outdoors during school days separately for summer and winter. They also reported their time participating in organised PA and sport activities during school days. Screen time assessment involved quantifying their time spent watching TV and using personal computers (PC) for entertainment purposes on school days. The four possible answers for all listed questions included “None” and “Less than one hour”, “One to two hours”, and “More than three hours” per day. Additionally, participants’ mothers were asked at all time points about their child’s mean sleep duration on school days. For the purpose of the present study, participants were labelled as “Meeting” and “Not meeting” optimal sleep duration for specific age category [18].

### Covariates

In this study, sex of participants and maternal education were employed as covariates. Maternal education, initially recorded in nine categories, was dichotomised into “non-university education” and “university degree” to maintain meaningful interpretation and allow comparison with other studies. Multiple imputation method, involving 10 iterations and 10 imputed datasets, was used to impute missing data for maternal education (*n* = 4). The imputation procedure was employed under the assumption that the data were missing completely at random.

### Statistical analysis

Descriptive statistics were calculated using the R software version 4.3.1 [19]. Latent transition analysis (LTA) was conducted using LatentGold version 6.0 (Statistical Innovations, Arlington, USA). LTA, a type of latent Markov model [20], was employed to analyse longitudinal data, identifying distinct typologies and transitions between them at different time points [21]. Activity-related behaviours served as input variables for the LTA models.

We tested models with 2 to 5 typologies to determine the optimal solution. Model fit was assessed using the Bayesian Information Criterion (BIC) and Akaike Information Criterion (AIC) [22], which evaluate the trade-off between model fit and complexity. Goodness of fit was also evaluated using the Vuong likelihood-ratio test, comparing models with *n* + 1 typologies to those with *n* typologies (*p* < 0.001 indicates that the *n* model is superior to the *n* + 1 typology model). Entropy was employed to assess the certainty of classifying individuals into distinct typologies based on their activity-related behaviour patterns, with values closer to 1 indicating a better fit. Additionally, we examined the size and meaning of the identified typologies, ensuring that minimum typology size was not less than 10% of the total sample size, averaged across all time points [21, 23, 24].

Due to a theoretical relationship and high bivariate residuals (BVR = 27.6) [25] between outdoor time during summer and winter, we included a direct effect between these two variables, to account for possible local dependencies and unexplainable variance accounted by the LTA model. To verify how demographic indicators were associated with the typology membership at the initial state and transition probabilities, we used bias-adjusted three-step LTA method [26]. Descriptive statistics for each typology were calculated as weighted proportions, using the posterior probability of assignment to a given typology as a weight for each observation.

### Patient and public involvement

Patients and the public were not involved in this study’s design, conduct, reporting or dissemination plans.

## Results

The sample (*n* = 380) was comprised of a 57% of girls and approximately one-third (35%) of participants’ mothers had a university degree. Based on the selection criteria, the best-fitting model revealed the optimum of four typologies across the three time points (Table 1). The response probabilities for each typology at baseline are presented in Table 2. The four typologies can be described as *Actives*, characterised by the highest proportion of time spent outdoors, time spent in organised PA and sports, and a low level of screen time (both in front of TV and PC); *Active screeners*, characterised by moderate levels of outdoor time and highest proportion of time spent in front of TV and PC; *Poor sleepers*, characterised by low proportion of meeting sleep recommendations and low time spent in front of PC. *Averages* was characterised by median levels of all activity-related behaviours except sleep, which was similar to other typologies except *Poor sleepers*. The most prevalent typology at the age of 11 years was *Averages* (65.1%), followed by *Active screeners* (21.0%), *Actives* (12.8%), and *Poor sleepers* (1.1%).

**Table 1.**
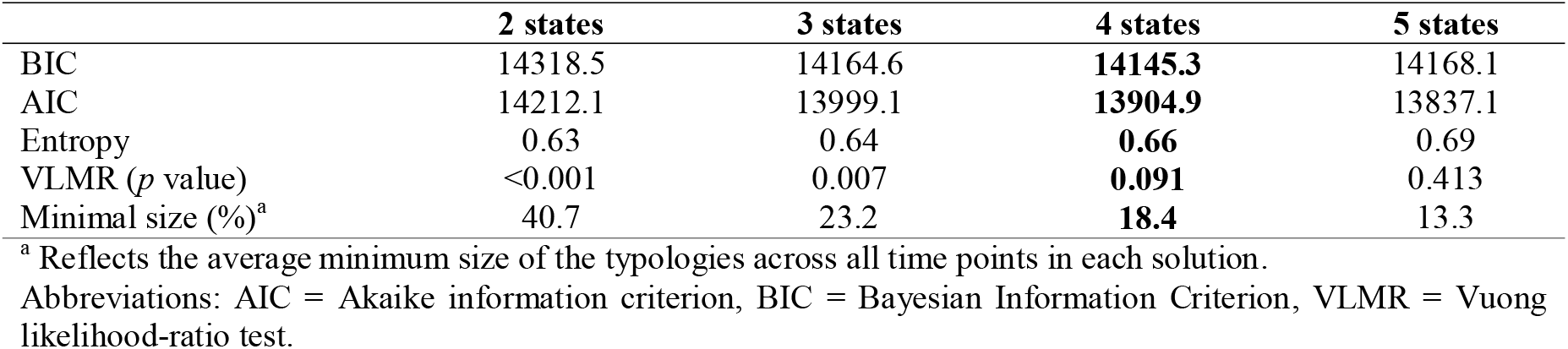
Statistical indicators for the transition models with 2-5 typologies.

**Table 2.** Response probabilities across the identified typologies at baseline.

|  | **Overall** | **Actives** | **Active screeners** | **Poor sleepers** | **Averages** |
| --- | --- | --- | --- | --- | --- |
|  | % | % | % | % | % |
| **Outdoor time during weekdays, average per day (summer)** |  |  |  |  |  |
| ≥3 hours | 42.7 | 72.0 | 54.9 | 35.7 | 27.7 |
| 1–2 hours | 42.0 | 25.6 | 38.0 | 47.7 | 48.4 |
| <1 hour | 14.0 | 2.4 | 6.8 | 15.3 | 21.6 |
| None | 1.3 | 0.1 | 0.3 | 1.2 | 2.4 |
| **Outdoor time during weekdays, average per day (winter)** |  |  |  |  |  |
| ≥3 hours | 9.9 | 22.6 | 14.2 | 1.9 | 6.2 |
| 1–2 hours | 44.5 | 57.8 | 54.4 | 28.0 | 42.0 |
| <1 hour | 41.2 | 19.2 | 30.0 | 59.7 | 47.3 |
| None | 4.4 | 0.5 | 1.3 | 10.4 | 4.6 |
| **Time spent in front of the PC, average per day** |  |  |  |  |  |
| ≥3 hours | 8.0 | 0.2 | 37.5 | 0.0 | 2.3 |
| 1–2 hours | 22.5 | 6.8 | 52.1 | 1.3 | 26.4 |
| <1 hour | 27.5 | 29.4 | 9.5 | 14.8 | 40.4 |
| None | 42.1 | 63.6 | 0.9 | 83.9 | 30.9 |
| **Time spent in front of the TV, average per day** |  |  |  |  |  |
| ≥3 hours | 12.2 | 15.7 | 28.1 | 3.1 | 7.9 |
| 1–2 hours | 49.2 | 56.9 | 57.1 | 34.8 | 49.1 |
| <1 hour | 32.2 | 24.8 | 13.9 | 47.0 | 36.5 |
| None | 6.4 | 2.6 | 0.8 | 15.1 | 6.5 |
| **Organised physical activity and sports participation, average per day** |  |  |  |  |  |
| ≥3 hours | 12.1 | 29.8 | 18.4 | 7.7 | 3.6 |
| 1–2 hours | 37.0 | 49.6 | 48.0 | 37.0 | 26.6 |
| <1 hour | 34.7 | 18.1 | 27.5 | 39.2 | 43.0 |
| None | 16.2 | 2.6 | 6.1 | 16.1 | 26.8 |
| **Sleep, average per day** |  |  |  |  |  |
| Meeting recommendation | 74.0 | 76.7 | 87.0 | 37.5 | 84.4 |
| Not meeting recommendation | 26.0 | 23.3 | 13.0 | 62.5 | 15.6 |

Sleep recommendations: 11 years old: 9–11 hours, 15 and 18 years old: 8–10 hours [18]
Abbreviations: PC = personal computer, TV = Television.

The transition probabilities indicated that at least 53.2% of individuals remained stable in their typology during the transition from 11 to 15 years (Figure 1). From the *Actives* (72.1% stability), 18.3% transitioned to *Active screeners* and 9.3% to *Poor sleepers*, there was minimal probability of transitioning to *Averages* (0.3%). From *Active screeners* (85.9% stability), a higher proportion of individuals transitioned to *Actives* (10.6%), while the rest transitioned to *Poor sleepers* (2.6%) and to *Averages* (1.0%). Most individuals from the *Poor sleepers* (53.2% stability) at 11 years transitioned to *Actives* (24.6%) and to *Averages* (20.5%), while only 1.8% transitioned to *Active screeners*. Moreover, a considerable proportion (18.2%) of individuals from the *Averages* (66.8% stability) transitioned to the *Actives*, while 9.0% and 6.0% transitioned to *Poor sleepers* and *Active screeners*, respectively. At 15 years, the distribution of the typologies was relatively comparable with 11 years, with the highest proportion of individuals belonging to *Averages* (44.0%), followed by *Active screeners* (24.3%), *Actives* (23.6%), and *Poor sleepers* (8.1%).

**Figure 1.**
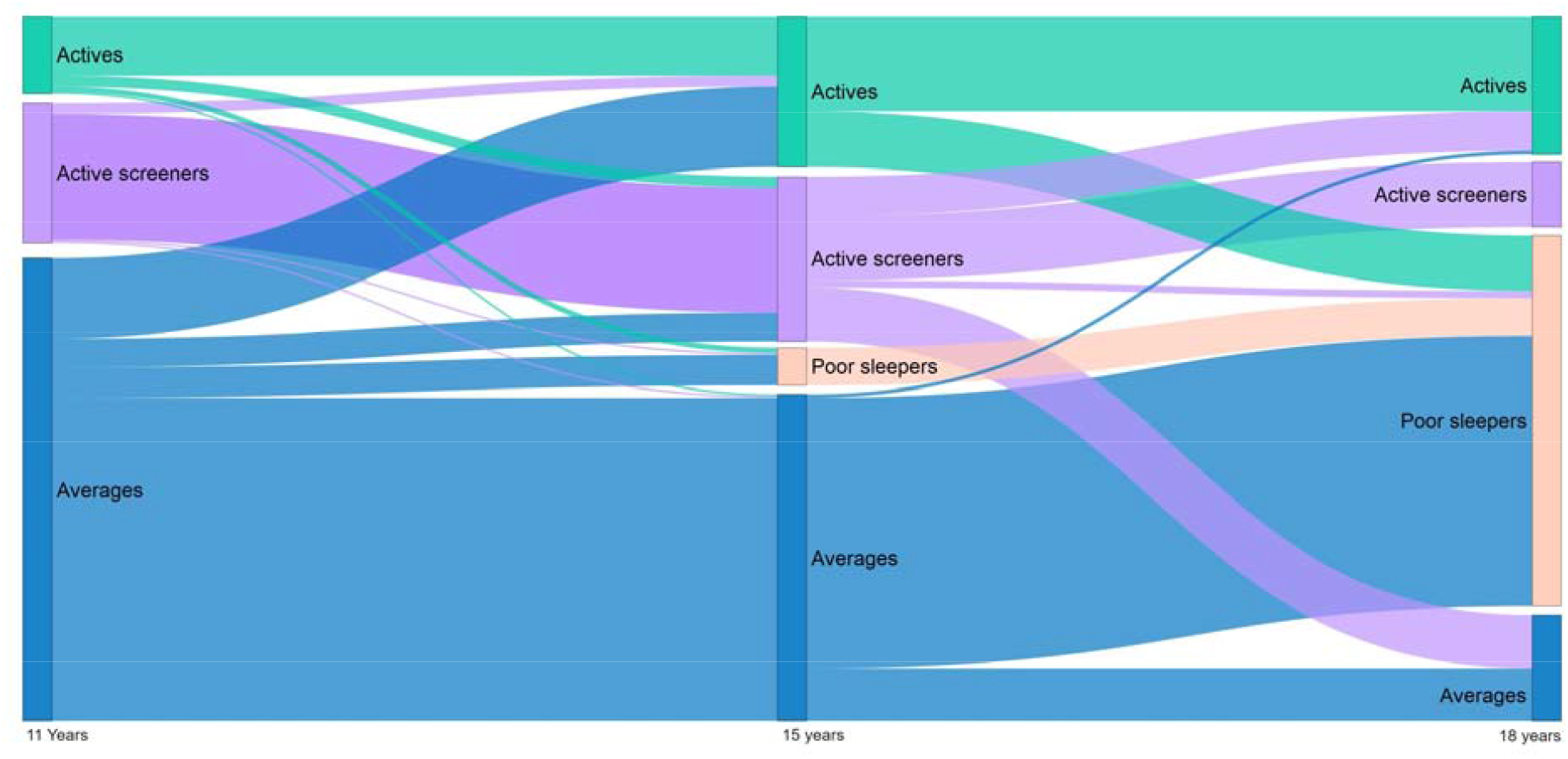
Illustration of transition probabilities across identified typologies (based on a hard assignment).

The observed transitions (Figure 1) between 15 and 18 years were notably different in comparison with the transitions from 11 to 15 years. From *Actives* (64.5% stability) most individuals transitioned to *Poor sleepers* (35.3%), while only 0.1% transitioned to each *Active screeners* and *Averages. Active screeners* (42.1% stability) transitioned mostly to the *Averages* (29.9%) and the *Actives* typologies (21.1%), while only 6.9% transitioned to the *Poor sleepers* typology. Minimal outflow from *Poor sleepers* was observed due to almost 99.5% stability in this typology, as only 0.2% transitioned to *Actives* and 0.1% transitioned to *Active screeners* and *Averages*. Most individuals transitioning from *Averages* (25.3% stability) transitioned to *Poor sleepers* (74.3%), only 0.3% transitioned to *Active screeners* and 0.1% to *Actives*. The final proportion of individuals in each typology at 18 years of age was represented by high dominance of *Poor sleepers* (50.8%) followed by *Actives* (20.4%) and *Averages* (18.4%), while only 10.4% of individuals were observed in the *Active screeners* typology.

The demographic characteristics of the identified typologies are presented in Table 3. At the initial time point, girls were more likely to be in the *Poor sleepers* typology in comparison with other typologies (*p* < 0.001). Sex was also associated with a probability of transitioning from one typology to another as boys were more likely to transition to *Averages* from other typologies at previous time point (*p* < 0.05 for all), while girls were more likely to transition to *Poor sleepers* from *Active screeners* (*p <* 0.001). Children of mothers with higher education were most prevalent in *Poor sleepers* (43%) and *Active screeners* (41%) (*p* = 0.038). The education of mothers was also associated with the transition probabilities (*p* = 0.002).

**Table 3.** Demographic indicators of the identified typologies at the initial time point.

|  | **Overall** | *Actives* | *Active Screeners* | *Poor Sleepers* | *Averages* | *p*-value<sup>1</sup> | *p*-value<sup>2</sup> |
| --- | --- | --- | --- | --- | --- | --- | --- |
|  | % | % | % | % | % |  |  |
| Girls, % | 56.6 | 65.7 | 20.1 | 76.7 | 66.2 | **<0.001** | **<0.001** |
| Mother university degree | 34.6 | 28.8 | 41.1 | 42.9 | 33.7 | **0.038** | **0.002** |

<sup>1</sup>Indicates associations between the indicators and typology membership
<sup>2</sup>Indicated associations between the indicator and transition probability between the typologies
Boldface values indicate significant difference between typologies at $p < 0.05$ .

## Discussion

Our study identified four distinct typologies of activity-related behaviours in children and adolescents based on their participation in organised PA and sports, screen time, outdoor time, and sleep. We also revealed that the proportion of typologies change during the transition from childhood and adolescence, and sex and maternal education status were associated with these changes. Specifically, there is a major shift in typology membership from 11 years to 18 years, with a decrease in the proportion of individuals in the *Active screeners* and *Averages* typologies and an increase in the proportion of individuals in the *Poor sleepers* typology. The most common transition was from the *Averages* to the *Poor Sleepers* typology.

The identified activity-related typologies differed in most of the indicator variables, except sleep as a high probability of optimal sleep duration was observed in three out of four identified typologies. However, increasing proportion of *Poor sleepers* highlights the importance of addressing sleep-related behaviours in childhood to prevent later sleep disturbance and associated health problems [6, 27]. An important finding from this study was that *Active screeners* were characterised by the combination of high screen time and high activity. As was previously demonstrated by Ferrar et al. [28], these behaviours can co-exist. The identification of the *Actives* typology suggests that there is a subgroup of individuals who engage in organised sports activities while also adhering to recommended sleep duration and having lower screen time compared to other typologies. A similar typology was observed in the study by Parker et al. [29], suggesting that this combination may be common in different contexts. However, this typology had a proportion below 24% during all three time points, and interventions should be targeted to ensure more children and adolescents have a similar combination of behaviours.

More than half of individuals remained stable in their typology during the transition from 11 to 15 years, however there were some notable transitions. The proportion of participants in the *Poor sleepers* typology was minimal at 11 years but rose in size with age, to become the largest typology at the age of 18 years. This suggests that there might be a shift towards unhealthy behaviours during adolescence [9, 30], which may be caused by changes in social dynamics or the potential influence of new responsibilities and commitments, such as earlier wake up times as a consequence of commuting to a secondary education or additional homework requirements. Conversely, the increased proportion of individuals in the *Actives* typology may indicate a potential increase in organised sports participation [31]. Approximately one in ten participants transitioned from the *Averages* to the *Actives* typology during this period, suggesting that some individuals may become more physically active as they transition into their teenage years. Future longitudinal studies may focus on these individuals helping to understand what makes them to improve their activity. *Averages* may benefit the most from intervention efforts, which could prevent the development of unhealthy activity patterns in later years.

A lower stability was observed throughout the transition from 15 to 18 years in comparison with the earlier period. While *Poor sleepers* and *Actives* remained relatively stable, *Active screeners* and *Averages* manifested strong outflow. With almost half of the proportion of the total sample at 15 years, three quarters of *Averages* transitioned to *Poor sleepers* by 18 years. These observations indicate that individuals with moderate levels of activity-related behaviours and optimal sleep duration may be inclined to unhealthy sleeping habits when they get older. On the other hand, more than 20% of *Active screeners* transitioned to *Actives*, as these individuals decreased their screen time. However, the *Actives* typology consisted mainly of individuals from the *Actives* and *Active screeners* typologies, suggesting that active individuals may maintain a higher amount of activity to early adulthood relative to others. It is necessary to note that our results are in contrast to previous research focusing on transitions of PA and SB typologies from adolescence to early adulthood (age 16–18) indicating over 80% of stability [29]. These differences might be accounted for by different indicator variables used in our analysis, such as the inclusion of sleep and outdoor time, but also to sample differences as person-oriented approaches may be sample-sensitive [32].

The association between demographic characteristics, such as sex and maternal education, with typology membership and transitions provides valuable insights into the influence of socio-demographic factors on activity-related behaviours in children and adolescents. For example, the study revealed that boys were more likely to be classified in the *Active screeners* typology compared to girls. This aligns with previous research suggesting that boys tend to engage in more screen-based activities while staying active compared to girls [14, 28]. Furthermore, our study found that individuals with mothers who had a university degree were more likely to be in the *Active screeners* typology compared to those with lower maternal education. These findings are in contrast to evidence that suggests a positive association between higher parental education and lower screen time [33]. However, our study focused on screen time for entertainment purposes on school days, inclusion of education-based and weekend screen time may provide different results. Additionally, both *Active screeners* and *Poor sleepers* had over 40% of highly educated mothers, but also the highest sex differences were observed between these two typologies suggesting that sex might be another important predictor of screen time. Nevertheless, it is necessary to interpret these findings cautiously, as these may be influenced by various contextual factors, such as parental modelling, which may play a different role at different stages of development of the children [33].

Our study is one of the first to examine activity-related typologies and their transitions from childhood to late adolescence. The use of a longitudinal design allowed for the examination of activity-related behaviour typologies and their transitions over time. Additionally, utilising previously under-researched activity-related behaviour variables (i.e., outdoor time and screen time) provided a comprehensive understanding of the lifestyle of children and adolescents.

Several limitations should also be acknowledged. One limitation is that the study relies on self-reported data, which may be subject to recall bias or social desirability bias. Also, we have used only data about behaviours during school days when there might be a significant variability of movement behaviours between school and weekend days [34, 35]. Additionally, we used cut-points for delivering optimal sleep duration to mitigate possible extreme values. Using a continuous sleep duration may provide different results. Furthermore, the generalizability of the findings may be limited as the study sample was drawn from a specific geographic region and may not be representative of broader populations. Conducting further research with larger and more diverse samples would help to strengthen the generalizability of these findings. Lastly, LTA was used to identify the typologies, which may introduce some degree of subjectivity in the classification process, as other studies may yield different results.

Future research could delve deeper into the specific factors (e.g. social and environmental) influencing the transitions between these typologies, allowing for a more nuanced understanding of the dynamics of activity-related behaviours during childhood and adolescence. This could lead to the development of targeted interventions that address the specific needs of different groups and explore mental and physical health consequences, ultimately promoting healthier activity-related behaviours and overall well-being.

## Conclusion

Four distinct typologies of activity-related behaviours in children and adolescents were identified, along with shifts in typologies over time, shedding light on the dynamic nature of these behaviours during a critical period of development. Most children remained in the same typology between ages 11-15, whereas there was more variability between 15-18 years. This highlights the significant shifts in lifestyle and behavioural patterns, particularly the increase in the proportion of individuals with poor sleeping patterns. These findings underscore the importance of implementing interventions that target multiple behavioural patterns and are oriented on individual characteristics to maintain and improve lifestyle quality.

## Declarations

### A funding statement

The CELSPAC-CZ study is supported by the RECETOX RI project financed by MEYS (LM2023069). This work is supported from the European Union’s Horizon 2020 research and innovation program under grant agreement No 857560. This publication reflects only the author’s view, and the European Commission is not responsible for any use that may be made of the information it contains. DJ is supported by student research grant (DSGC-2021-0022), which was funded by the OP RDE project “Improving schematics of Doctoral student grant competition and their pilot implementation”, with Reg. No.: CZ.02.2.69/0.0/0.0/19_073/0016713 and from the Palacký University Olomouc internal grant (IGA_FTK_2023_001). AG is supported by the Czech Science Foundation (22-02392S). LAr is supported by an Australian Research Council Discovery Early Career Researcher Award (DE220100847).

### A competing interests statement

The authors declare that they have no competing interests.

## Author contributions

DJ drafted the initial manuscript, conceptualised the study, and conducted the analyses; AG, AMCA, AT, and LAr conceptualised and designed the study, contributed to writing and critically revied and edited the manuscript; LAn and PP were responsible for the data curation and critically revised the manuscript; All authors have read and approved the final version of the manuscript, and agree with the order of presentation of the authors.

## Availability of data and materials

The data that support the findings of this study are available from www.elspac.cz by Research Centre for Toxic Compounds in the Environment (RECETOX) but restrictions apply to the availability of these data, which were used under license for the current study, and so are not publicly available.

## Notes

### Competing Interest Statement

The authors have declared no competing interest.

### Author Declarations

The study was approved by the ELSPAC-CZ Law and Ethics Committee and local research ethics committees. The secondary use of all ELSPAC-CZ study data was approved by the (C)ELSPAC Ethics Committee (Ref. No. ELSPAC/EK/1/2014, date 09/17/2014).

